# Infliximab is associated with attenuated immunogenicity to BNT162b2 and ChAdOx1 nCoV-19 SARS-CoV-2 vaccines

**DOI:** 10.1101/2021.03.25.21254335

**Authors:** Nicholas A Kennedy, Simeng Lin, James R Goodhand, Neil Chanchlani, Ben Hamilton, Claire Bewshea, Rachel Nice, Desmond Chee, JR Fraser Cummings, Aileen Fraser, Peter M Irving, Nikolaos Kamperidis, Klaartje B Kok, Christopher A Lamb, Jonathan Macdonald, Shameer J Mehta, Richard CG Pollok, Tim Raine, Philip J Smith, Ajay M Verma, Timothy J McDonald, Shaji Sebastian, Charlie W Lees, Nick Powell, Tariq Ahmad, Contributors to the CLARITY IBD study

**Affiliations:** Department of Gastroenterology, Royal Devon and Exeter NHS Foundation Trust, Exeter, UK; Exeter Inflammatory Bowel Disease and Pharmacogenetics Research Group, University of Exeter, UK; Department of Biochemistry, Exeter Clinical Laboratory International, Royal Devon and Exeter NHS Foundation Trust, Exeter, UK; Department of Gastroenterology, University Hospital Southampton NHS Foundation Trust, Southampton, UK; Department of Gastroenterology, University Hospitals Bristol NHS Foundation Trust, Bristol, UK; Department of Gastroenterology, Guy’s and St Thomas’ NHS Foundation Trust, London, UK; School of Immunology & Microbial Sciences, King’s College London, London, UK; Department of Gastroenterology, St Marks Hospital and Academic Institute, London, UK; Centre for Immunobiology, Blizard Institute, Barts and the London School of Medicine, Queen Mary University of London, London, UK; Department of Gastroenterology, Royal London Hospital, Barts Health NHS Trust, London, UK; Department of Gastroenterology, Newcastle upon Tyne Hospitals NHS Foundation Trust, Newcastle upon Tyne, UK; Translational & Clinical Research Institute, Faculty of Medical Sciences, Newcastle University, Newcastle upon Tyne, UK; Department of Gastroenterology, Queen Elizabeth University Hospital, NHS Greater Glasgow and Clyde, Glasgow, UK; School of Medicine, Dentistry and Nursing, University of Glasgow, Glasgow, UK; Department of Gastroenterology, University College London Hospitals NHS Foundation Trust, London, UK; Department of Gastroenterology, St George’s University Hospital NHS Foundation Trust, London, UK; Institute for Infection & Immunity, St George’s, University of London, London, UK; Department of Gastroenterology, Addenbrooke’s Hospital, Cambridge University Hospitals NHS Foundation Trust, Cambridge, UK; Department of Gastroenterology, Royal Liverpool Hospital, Liverpool University Hospitals NHS Foundation Trust, Liverpool, UK; Department of Gastroenterology, Kettering General Hospital, Kettering, UK; IBD Unit, Department of Gastroenterology, Hull University Teaching Hospitals NHS Trust, Hull, UK; Hull York Medical School, University of Hull, Hull, UK; Department of Gastroenterology, Western General Hospital, NHS Lothian, Edinburgh, UK; Institute of Genetic and Molecular Medicine, University of Edinburgh, Edinburgh, UK; Department of Gastroenterology, Western General Hospital, Edinburgh, UK; Department of Gastroenterology, Imperial College Healthcare NHS Trust, London, UK; Department of Metabolism, Digestion and Reproduction, Imperial College London, London, UK

**Author notes:** **Address for correspondence:** Prof Tariq Ahmad, Exeter Inflammatory Bowel Disease and Pharmacogenetics Research Group, RILD building, Barrack Road, Exeter. EX2 5DW, UK. denotes equal contribution (Nicholas A Kennedy, Simeng Lin, James R Goodhand, Nick Powell, Tariq Ahmad).

**Keywords:** SARS-CoV-2, immune-mediated inflammatory diseases, inflammatory bowel disease, anti-TNF therapy, infliximab, vedolizumab, immunosuppressant, vaccine, ChAdOx1 nCoV-19, BNT162b2, CLARITY

## Abstract

**Background:** Delayed second-dose SARS-CoV-2 vaccination trades maximal effectiveness for a lower level of immunity across more of the population. We investigated whether patients with inflammatory bowel disease treated with infliximab have attenuated serological responses to a single-dose of a SARS-CoV-2 vaccine.

**Methods:** Antibody responses and seroconversion rates in infliximab-treated patients (n=865) were compared to a cohort treated with vedolizumab (n=428), a gut-selective anti-integrin α4β7 monoclonal antibody. Our primary outcome was anti-SARS-CoV-2 spike (S) antibody concentrations 3-10 weeks after vaccination in patients without evidence of prior infection. Secondary outcomes were seroconversion rates, and antibody responses following past infection or a second dose of the BNT162b2 vaccine.

**Findings:** Geometric mean [SD] anti-SARS-CoV-2 antibody concentrations were lower in patients treated with infliximab than vedolizumab, following BNT162b2 (6.0 U/mL [5.9] vs 28.8 U/mL [5.4] P<0.0001) and ChAdOx1 nCoV-19 (4.7 U/mL [4.9]) vs 13.8 U/mL [5.9] P<0.0001) vaccines. In our multivariable models, antibody concentrations were lower in infliximab-compared to vedolizumab-treated patients who received the BNT162b2 (fold change [FC] 0.29 [95% CI 0.21, 0.40], p<0.0001) and ChAdOx1 nCoV-19 (FC 0.39 [95% CI 0.30, 0.51], p<0.0001) vaccines. In both models, age ≥ 60 years, immunomodulator use, Crohn’s disease, and smoking were associated with lower, whilst non-white ethnicity was associated with higher, anti-SARS-CoV-2 antibody concentrations. Seroconversion rates after a single-dose of either vaccine were higher in patients with prior SARS-CoV-2 infection and after two doses of BNT162b2 vaccine.

**Interpretation:** Infliximab is associated with attenuated immunogenicity to a single-dose of the BNT162b2 and ChAdOx1 nCoV-19 SARS-CoV-2 vaccines. Vaccination after SARS-CoV-2 infection, or a second dose of vaccine, led to seroconversion in most patients. Delayed second dosing should be avoided in patients treated with infliximab.

**Funding:** Royal Devon and Exeter and Hull University Hospital Foundation NHS Trusts. Unrestricted educational grants: F. Hoffmann-La Roche AG (Switzerland), Biogen GmbH (Switzerland), Celltrion Healthcare (South Korea) and Galapagos NV (Belgium).

**Research in context:** *Evidence before this study:* Faced with further surges of SARS-CoV-2 infection, a growing number of countries, including the UK, have opted to delay second vaccine doses for all people. This strategy trades maximal effectiveness against a lower level of protective immunity across more of the at-risk population. We have previously shown that seroprevalence, seroconversion in PCR-confirmed cases, and the magnitude of anti-SARS-CoV-2 antibodies following SARS-CoV-2 infection are reduced in infliximab-compared with vedolizumab-treated patients. Whether single-doses of vaccines are effective in patients treated with anti-TNF therapies is unknown. We searched PubMed from 25 November 2019 to 23 March 2021 with the terms “anti-tumour necrosis factor” or “anti-integrin” or “infliximab” or “adalimumab” or “vedolizumab” or “biological therapy” or “biologic therapy” AND “SARS-CoV-2” or “coronavirus” or “COVID-19” or AND “seroprevalence” or “seroconversion” or “antibody” or “antibody response” or “magnitude” or “immunogenicity” AND “vaccine” or “vaccination” or “immunisation” or “immunization” or “ChAdOx1 nCoV-19” or “BNT162b2” or “mRNA-1273”, without restriction on language. Serological responses to SARS-CoV-2 vaccines have been reported in registration trials and small observational cohorts of healthy volunteers. Two small studies, including one unpublished preprint, found that COVID-19 vaccine immunogenicity rates were lower in transplant recipients and patients with malignancy receiving immunosuppressive therapy, and fewer patients treated with potent immunosuppressants seroconverted than healthy controls. No studies have assessed the effect of anti-TNF therapy on immunogenicity following SARS-CoV-2 vaccination.

*Added value of this study:* To test if anti-TNF drugs attenuate serological responses to primary SARS-CoV-2 vaccines, we analysed anti-SARS-CoV-2 spike (S) antibody concentrations and seroconversion rates in 1293 patients with inflammatory bowel disease who received primary vaccinations with either the ChAdOx1 nCoV-19 or BNT162b2 vaccines. 865 were treated with the anti-TNF drug infliximab and outcomes were compared to a reference cohort of 428 patients treated with vedolizumab, a gut selective anti-integrin α4β7 monoclonal antibody that is not associated with impaired systemic immune responses. Anti-SARS-CoV-2 antibody levels and rates of seroconversion were lower following primary vaccination with both the BNT162b2 and ChAdOx1 nCoV-19 vaccines in patients with IBD treated with infliximab compared to vedolizumab. Older age, immunomodulator use, Crohn’s disease (versus ulcerative colitis or inflammatory bowel disease unclassified), and current smoking were associated with lower anti-SARS-CoV-2 antibody concentrations, irrespective of vaccine type. Non-white ethnicity was associated with higher anti-SARS-CoV-2 (S) antibody concentrations following primary vaccination with both vaccines. Antibody concentrations and seroconversion rates were higher in patients with past SARS-CoV-2 infection prior to a single-dose of either vaccine, and after 2 doses of the BNT162b2 vaccine.

*Implications of the available evidence:* Our findings have important implications for patients treated with anti-TNF therapy, particularly for those also treated with an immunomodulator. Poor antibody responses to a single-dose of vaccine exposes these patients to a potential increased risk of SARS-CoV-2 infection. However, higher rates of seroconversion in patients with two exposures to SARS-CoV-2 antigen, even in the presence of TNF blockade, suggest that all patients receiving these drugs should be prioritized for optimally timed second doses. Until patients receive a second vaccine dose, they should consider that they are not protected from SARS-CoV-2 infection and continue to practice enhanced physical distancing and shielding if appropriate. Even after two antigen exposures, a small subset of patients failed to mount an antibody response. Antibody testing and adapted vaccine schedules should be considered to protect these at-risk patients.

## Introduction

Limited SARS-CoV-2 vaccine supplies and pressure on critical care services have forced governments to prioritise primary vaccination to vulnerable groups. In the United Kingdom, second vaccine doses have also been delayed, trading maximal effectiveness for a lower level of protective immunity across a greater proportion of the most at-risk population.^1^ Consequently, more than half of the adult population have received a single-dose of either the RNA vaccine, BNT162b2 (Pfizer/BioNTech) or the adenovirus-vector vaccine, ChAdOx1 nCoV-19 (Oxford/AstraZeneca). Faced with further surges of SARS-CoV-2 infection, a growing number of other countries have also opted to delay second vaccine doses.^2,3^

The inflammatory bowel diseases (IBD), Crohn’s disease and ulcerative colitis (UC) are chronic immune-mediated inflammatory diseases (IMIDs) that affect about 1% of the UK population.^4,5^ Treatment typically requires immunosuppression with immunomodulators (azathioprine, mercaptopurine, and methotrexate) and/or biological therapies that target disease relevant cytokines or the immune cells that produce them. Anti-tumour necrosis factor (TNF) drugs, such as infliximab and adalimumab, are the most frequently prescribed biopharmaceuticals used in the treatment of IMIDs. These drugs impair immunogenicity following pneumococcal,^6^ influenza,^7^ and hepatitis B^8^ vaccinations and increase the risk of serious infection, most notably with respiratory pathogens.^9^ Conversely, vedolizumab, a gut-selective anti-integrin α4β7 monoclonal antibody is not associated with increased susceptibility to systemic infection or attenuated serological responses to vaccination.^10^

We have recently reported that seroprevalence, seroconversion in PCR-confirmed cases, and the magnitude of anti-SARS-CoV-2 antibodies following SARS-CoV-2 infection are reduced in infliximab-compared with vedolizumab-treated patients.^11^ We hypothesised that, following at least a single-dose with BNT162b2 or ChAdOx1 nCoV-19 vaccine, serological responses would be similarly impaired in patients treated with infliximab compared to vedolizumab arguing against delaying second doses in these patients.

We aimed to define, in patients with IBD who had received a COVID-19 vaccination, whether biologic class and concomitant use of an immunomodulator impact:

i. anti-SARS-CoV-2 spike (S) antibody levels
ii. rates of seroconversion
iii. antibody responses in patients who had previously been infected with SARS-CoV-2 or who had two doses of vaccine

## Methods

### Study design and participants

impaCt of bioLogic therApy on saRs-cov-2 Infection and immuniTY (CLARITY) IBD is a UK wide, multicentre, prospective observational cohort study investigating the impact of infliximab and vedolizumab and/or concomitant immunomodulators (azathioprine, mercaptopurine, and methotrexate) on SARS-CoV-2 acquisition, illness, and immunity in patients with IBD.

Study methods have been described in detail previously.^11^ In brief, consecutive patients were recruited at the time of attendance at infusion units from 92 National Health Service (NHS) hospitals across the UK between 22^nd^ September 2020 and 23^rd^ December 2020 (Supplementary pp 2 - 17). The eligibility criteria were age 5 years and over, a diagnosis of IBD, and current treatment with infliximab or vedolizumab for 6 weeks or more, with at least one dose of drug received in the previous 16 weeks. Patients were excluded if they had participated in a SARS-CoV-2 vaccine trial.

Follow-up visits were timed to coincide with biologic infusions and occurred approximately eight-weekly. Here, we report vaccine-induced antibody responses at first study visit after primary vaccination, and where possible, after two doses. Participants were eligible for inclusion in our vaccine immunogenicity analysis if they had had a SARS-CoV-2 antibody test within the first ten weeks after their primary vaccination with any of the available SARS-CoV-2 vaccines.

The Surrey Borders Research Ethics committee approved the study (REC reference: 20/HRA/3114) in September 2020. Patients were included after providing informed, written consent. The sponsor was the Royal Devon and Exeter NHS Foundation Trust. The protocol is available online at https://www.clarityibd.org. The study was registered with the ISRCTN registry, ISRCTN45176516.

### Procedures

Variables recorded by participants were demographics (age, sex, ethnicity, comorbidities, height and weight, smoking status, and postcode), IBD disease activity (PRO2), SARS-CoV-2 symptoms aligned to the COVID-19 symptoms study (symptoms, previous testing, and hospital admissions for COVID-19), and vaccine uptake (type and date of primary vaccination). Study sites completed data relating to IBD history (age at diagnosis, disease duration, and phenotype according to the Montreal classifications, previous surgeries, and duration of current biologic and immunomodulator therapy).^11^ We linked our data by NHS number or Community Health Index to Public Health England, Scotland, and Wales who archive dates and results of all SARS-CoV-2 PCR tests undertaken. Data were entered electronically into a purpose-designed REDCap database hosted at the Royal Devon and Exeter NHS Foundation Trust.^12^ Participants without access to the internet or electronic device completed their questionnaires on paper case record forms that were subsequently entered by local research teams.

Laboratory analyses were performed at the Academic Department of Blood Sciences at the Royal Devon and Exeter NHS Foundation Trust. To determine antibody responses specific to vaccination we used the Roche Elecsys Anti-SARS-CoV-2 spike (S) immunoassay^13^ alongside the nucleocapsid (N) immunoassay.^14^ This double sandwich electrochemiluminescence immunoassay uses a recombinant protein of the receptor binding domain on the spike protein as an antigen for the determination of antibodies against SARS-CoV-2. Sample electrochemiluminescence signals are compared to an internal calibration curve and quantitative values are reported as units (U)/mL.

In-house assay validation experiments demonstrated:

i. The intra-assay and inter-assay coefficient of variation were 1.3% and 5.6%, respectively
ii. Anti-SARS-CoV-2 (S) antibodies were stable in uncentrifuged blood and serum at ambient temperature for up to seven days permitting postal transport
iii. No effect was observed on recovery of anti-SARS-CoV-2 (S) antibodies following four freeze/thaw cycles
iv. No analytical interference was observed for the detection of anti-SARS-CoV-2 (S) with infliximab or vedolizumab up to 10,000 mg/L and 60,000 mg/L, respectively, or with anti-drug antibodies to infliximab or vedolizumab up to 400 AU/mL and 38 AU/mL, respectively (data not shown).

At entry to CLARITY IBD and at follow-up visits, all patients were tested for previous SARS-CoV-2 infection using the Roche Elecsys anti-SARS-CoV-2 (N) immunoassay. Because antibody responses are impaired following PCR-confirmed natural infection we set a threshold of 0.25 times the cut-off index (COI) at or above which patients were deemed to have had prior infection.^11^ We defined a second threshold of 0.12 times the COI, below which patients were deemed to have no evidence of prior infection. Patients with a PCR test confirming SARS-CoV-2 infection at any time prior to vaccination were deemed to have evidence of past infection irrespective of any antibody test result.

### Outcomes

Our primary outcome was anti-SARS-CoV-2 anti-spike (S) protein receptor-binding protein antibodies three to ten weeks after primary vaccination.

Secondary outcomes were:

i. proportion of participants with seroconversion, defined by a threshold which has been associated with pseudoneutralisation in vitro.
ii. antibody concentrations and seroconversion in patients with PCR or serological evidence of past SARS-CoV-2 infection at, or prior, to the post-vaccination serum sample.
iii. antibody concentrations and seroconversion after two doses of vaccine.

### Statistical Analysis

The sample size for CLARITY IBD was based on the number of participants required to demonstrate a difference in the impact of infliximab and vedolizumab on seroprevalence and seroconversion following SARS-CoV-2 infection, with an estimated background seroprevalence of 0.05. We calculated that a sample of 6970 patients would provide 80% power to detect differences in the seroprevalence of anti-SARS-CoV-2 antibodies in infliximab-compared with vedolizumab-treated patients, whilst controlling for immunomodulator status at the 0.05 significance level. We stored and then analysed all serum samples as soon as the Roche Elecsys anti-SARS-CoV-2 (S) immunoassay was established in our laboratory.

Statistical analyses were undertaken in R 4.0.4 (R Foundation for Statistical Computing, Vienna, Austria). All tests were two tailed and p-values <0.05 were considered significant. We included patients with missing clinical data in analyses for which they had data and have specified the denominator for each variable. Anti-S antibody concentrations are reported as geometric means and standard deviations. Other continuous data are reported as median and interquartile range, and discrete data as numbers and percentages, unless otherwise stated.

Univariable analyses, using t-tests of log-transformed anti-SARS-CoV-2 (S) antibody concentration and Spearman’s rank correlation coefficients, were used to identify demographic, disease, vaccine, and treatment-related factors associated with the concentration of anti-SARS-CoV-2 (S) antibodies. To test our primary outcome, we used multivariable linear regression models to identify factors independently associated with log anti-SARS-CoV-2 (S) levels. A priori, we included age, ethnicity, biologic medication, and immunomodulator use. No stepwise regression was performed. Results are presented after exponentiation, so that the coefficients of the model correspond to the fold change associated with each binary covariate. For age, a cut-off was chosen based on graphical inspection of the relationship between age and anti-SARS-CoV-2 (S) antibody concentrations. We also report the proportions of patients who seroconverted following vaccination. Seroconversion was defined by an optimized cut-off of 15 U/mL, with a positive predictive value of 99.1%, based on the correlation between receptor-binding domain antibodies and the cPass SARS-CoV-2 surrogate virus neutralisation test (internal data, Roche Diagnostics GmbH, Germany).^15,16^ We conducted sensitivity analyses to compare antibody responses stratified by participants with serological or PCR evidence of SARS-CoV-2 infection at any time prior to vaccination and in those who had received 2 doses of vaccine.

### Role of the funding source

CLARITY IBD is an investigator-led, UK National Institute for Health Research COVID-19 urgent public health study, funded by the Royal Devon and Exeter NHS Foundation Trust, Hull University Teaching Hospital NHS Trust, and by unrestricted educational grants from F. Hoffmann-La Roche AG (Switzerland), Biogen GmbH (Switzerland), Celltrion Healthcare (South Korea) and Galapagos NV (Belgium). None of our funding bodies had any role in study design, data collection or analysis, writing, or decision to submit for publication.

## Results

Between September 22^nd^ 2020 and December 23^rd^ 2020, 7226 patients were recruited to the CLARITY study from 92 UK hospitals.^11^ For the primary immunogenicity analyses we included 865 infliximab- and 428 vedolizumab-treated participants without evidence of prior SARS-CoV-2 infection, who had received uninterrupted biologic therapy since recruitment and had an antibody test between 21 and 70 days after primary vaccination. Participant characteristics are shown in Table 1.

**Table 1:** Baseline characteristics of participants who had anti-SARS-CoV-2 spike antibodies measured 3 to 10 weeks following primary vaccination against SARS-CoV-2.

| Variable |  | Infliximab | Vedolizumab | Overall | p |
| --- | --- | --- | --- | --- | --- |
| Vaccine | BNT162b2 | 44.7% (387/865) | 47.2% (202/428) | 45.6% (589/1293) | 0.41 |
|  | ChAdOx1 nCoV-19 | 55.3% (478/865) | 52.8% (226/428) | 54.4% (704/1293) |  |
| Age (years) |  | 41.4 (31.5 - 54.8) | 49.6 (37.1 - 63.8) | 43.8 (32.8 - 57.6) | <0.0001 |
| Sex | Female | 50.3% (434/863) | 47.1% (200/425) | 49.2% (634/1288) | 0.19 |
|  | Male | 49.7% (429/863) | 52.7% (224/425) | 50.7% (653/1288) |  |
|  | Intersex | 0.0% (0/863) | 0.0% (0/425) | 0.0% (0/1288) |  |
|  | Prefer not to say | 0.0% (0/863) | 0.2% (1/425) | 0.1% (1/1288) |  |
| Ethnicity | White | 91.8% (791/862) | 89.9% (381/424) | 91.1% (1172/1286) | 0.62 |
|  | Asian | 5.3% (46/862) | 7.5% (32/424) | 6.1% (78/1286) |  |
|  | Mixed | 1.9% (16/862) | 1.9% (8/424) | 1.9% (24/1286) |  |
|  | Black | 0.7% (6/862) | 0.5% (2/424) | 0.6% (8/1286) |  |
|  | Other | 0.3% (3/862) | 0.2% (1/424) | 0.3% (4/1286) |  |
| Diagnosis | Crohn's disease | 65.4% (566/865) | 40.7% (174/428) | 57.2% (740/1293) | 0.00050 |
|  | Ulcerative colitis or IBD-unclassified | 34.6% (299/865) | 59.3% (254/428) | 42.8% (553/1293) |  |
| Duration of IBD (years) |  | 8.0 (4.0 - 16.0) | 10.0 (5.0 - 17.8) | 9.0 (4.0 - 16.0) | 0.0040 |
| Age at IBD diagnosis (years) |  | 28.8 (21.6 - 41.8) | 34.0 (23.3 - 47.6) | 30.3 (21.9 - 43.7) | <0.0001 |
| Immunomodulator |  | 61.6% (533/865) | 22.0% (94/427) | 48.5% (627/1292) | <0.0001 |
| 5-ASA |  | 23.0% (199/865) | 31.6% (135/427) | 25.9% (334/1292) | 0.0012 |
| Steroids |  | 3.0% (26/865) | 8.4% (36/427) | 4.8% (62/1292) | <0.0001 |
| BMI (kg/m<sup>2</sup>) |  | 25.9 (22.8 - 30.6) | 26.1 (23.1 - 30.1) | 26.0 (22.9 - 30.4) | 0.75 |
| Heart disease |  | 3.6% (31/865) | 6.5% (28/428) | 4.6% (59/1293) | 0.023 |
| Diabetes |  | 3.8% (33/865) | 7.5% (32/428) | 5.0% (65/1293) | 0.0065 |
| Lung disease |  | 13.5% (117/865) | 18.2% (78/428) | 15.1% (195/1293) | 0.032 |
| Kidney disease |  | 1.2% (10/865) | 2.1% (9/428) | 1.5% (19/1293) | 0.22 |
| Cancer |  | 0.5% (4/865) | 2.1% (9/428) | 1.0% (13/1293) | 0.013 |
| Smoker | Yes | 9.7% (84/862) | 5.4% (23/425) | 8.3% (107/1287) | 0.0010 |
|  | Not currently | 32.0% (276/862) | 41.6% (177/425) | 35.2% (453/1287) |  |
|  | Never | 58.2% (502/862) | 52.9% (225/425) | 56.5% (727/1287) |  |
| Exposure to documented cases of COVID-19 |  | 9.4% (81/862) | 8.7% (37/425) | 9.2% (118/1287) | 0.76 |
| Income deprivation score |  | 0.086 (0.052 - 0.151) | 0.084 (0.054 - 0.141) | 0.086 (0.052 - 0.147) | 0.94 |
| Active disease (PRO2) |  | 4.9% (41/831) | 11.4% (46/405) | 7.0% (87/1236) | <0.0001 |

**Abbreviations:** IBD = inflammatory bowel disease; 5-ASA = 5-aminosalicylic acid; BMI = Body Mass Index; PRO2 = IBD disease activity. Values presented are median (interquartile range) or percentage (numerator/denominator). P values represent the results of a Mann Whitney U, Kruskal Wallis or Fisher's exact test.

### Anti-SARS-CoV-2 (S) antibody level following primary COVID-19 vaccine

Geometric mean [geometric SD] anti-SARS-CoV-2 (S) antibody concentrations were lower in patients treated with infliximab than vedolizumab, following both the BNT162b2 (6.0 U/mL [5.9] vs 28.8 U/mL [5.4] P<0.0001) and ChAdOx1 nCoV-19 (4.7 U/mL [4.9] vs 13.8 U/mL [5.9] P<0.0001) vaccines (Figure 1). Amongst infliximab-treated patients, the geometric mean [geometric SD] anti-SARS-CoV-2 (S) antibody concentrations were also lower in patients treated with a concomitant immunomodulator. Additional univariable analyses are shown in Table 2.

**Table 2:** Univariable associations with anti-SARS-CoV-2 spike antibodies, stratified by vaccine type.

| Variable |  | BNT162b2 |  | ChAdOx1 nCoV-19 |  |
| --- | --- | --- | --- | --- | --- |
|  |  | Value | p | Value | p |
| Biologic treatment | Infliximab | 6.0 (5.9) | <0.0001 | 4.7 (4.9) | <0.0001 |
|  | Vedolizumab | 28.8 (5.4) |  | 13.8 (5.9) |  |
| Immunomodulator in infliximab-treated participants | No | 9.7 (4.7) | <0.0001 | 5.7 (5.1) | 0.045 |
|  | Yes | 4.4 (6.3) |  | 4.2 (4.7) |  |
| Immunomodulator in vedolizumab-treated participants | No | 32.4 (5.2) | 0.052 | 15.6 (6.0) | 0.082 |
|  | Yes | 16.7 (6.3) |  | 10.0 (5.5) |  |
| Age (years) |  | rho = -0.22 | <0.0001 | rho = -0.15 | <0.0001 |
| Sex | Female | 9.4 (7.0) | 0.092 | 6.6 (5.5) | 0.83 |
|  | Male | 10.9 (6.3) |  | 6.8 (5.7) |  |
| Ethnicity | White | 9.4 (6.6) | 0.037 | 6.2 (5.6) | 0.0051 |
|  | Asian | 20.9 (7.3) |  | 16.1 (5.2) |  |
|  | Mixed | 25.7 (6.7) |  | 13.7 (5.3) |  |
|  | Black | 12.5 (1.6) |  | 19.4 (2.2) |  |
|  | Other | 22.9 (3.7) |  | 5.7 (3.1) |  |
| Diagnosis | Crohn's disease | 7.3 (6.4) | <0.0001 | 5.6 (5.6) | 0.0014 |
|  | Ulcerative colitis or IBD-unclassified | 15.6 (6.5) |  | 8.5 (5.5) |  |
| Duration of IBD (years) |  | rho = -0.16 | <0.0001 | rho = -0.12 | 0.0013 |
| Age at IBD diagnosis (years) |  | rho = -0.13 | 0.0021 | rho = -0.04 | 0.25 |
| 5-ASA | No | 9.8 (6.6) | 0.40 | 6.7 (5.5) | 0.93 |
|  | Yes | 11.5 (7.1) |  | 6.6 (5.9) |  |
| Steroids | No | 10.2 (6.7) | 0.90 | 6.8 (5.5) | 0.12 |
|  | Yes | 10.7 (7.3) |  | 4.1 (6.7) |  |
| BMI (kg/m<sup>2</sup>) |  | rho = -0.08 | 0.068 | rho = -0.01 | 0.81 |
| Heart disease | No | 10.3 (6.7) | 0.65 | 6.9 (5.6) | 0.010 |
|  | Yes | 8.7 (7.0) |  | 2.8 (5.2) |  |
| Diabetes | No | 10.7 (6.7) | 0.0028 | 6.8 (5.6) | 0.066 |
|  | Yes | 4.1 (4.6) |  | 4.0 (5.2) |  |
| Lung disease | No | 10.1 (6.9) | 0.70 | 6.9 (5.5) | 0.31 |
|  | Yes | 10.9 (5.7) |  | 5.7 (6.1) |  |
| Kidney disease | No | 10.2 (6.6) | 0.60 | 6.7 (5.5) | 0.66 |
|  | Yes | 15.6 (10.4) |  | 4.7 (12.4) |  |
| Cancer | No | 10.4 (6.6) | 0.13 | 6.7 (5.6) | 0.069 |
|  | Yes | 2.0 (9.2) |  | 2.3 (3.6) |  |
| Smoking | Yes | 4.7 (7.1) | 0.0077 | 3.4 (4.8) | 0.00077 |
|  | Not currently | 9.4 (6.6) |  | 6.1 (5.4) |  |
|  | Never | 11.8 (6.5) |  | 8.0 (5.7) |  |
| Exposure to documented cases of COVID-19 | No | 10.3 (6.7) | 0.87 | 6.6 (5.5) | 0.53 |
|  | Yes | 9.8 (6.8) |  | 7.8 (6.1) |  |
| Income deprivation score |  | rho = 0.01 | 0.75 | rho = 0.02 | 0.65 |
| Active disease (PRO2) | No | 10.1 (6.5) | 0.32 | 6.6 (5.4) | 0.51 |
|  | Yes | 14.0 (7.6) |  | 8.1 (7.0) |  |

**Abbreviations:** IBD = inflammatory bowel disease; 5-ASA = 5-aminosalicylic acid; VAS = visual analogue scale. Values presented are geometric mean antibody concentration (geometric standard deviation) or Spearman's rho. P values represent the results of an unpaired t test or test of Spearman's rho.

**Figure 1:**
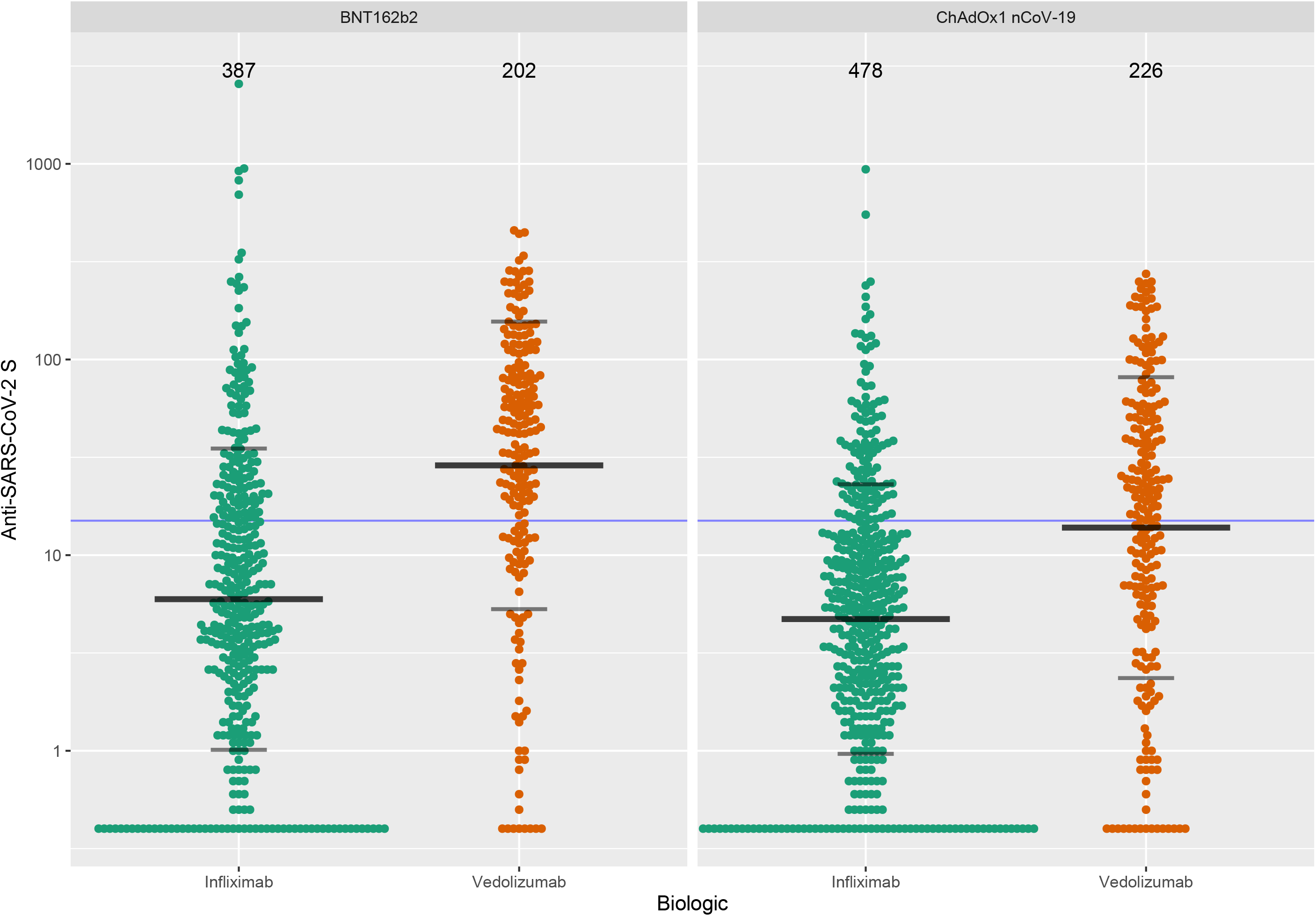
Anti-SARS-CoV-2 spike antibody concentration stratified by biologic therapy (infliximab vs vedolizumab) and type of vaccine. The wider bar represents the geometric mean, while the narrower bars are drawn one geometric standard deviation either side of the geometric mean. The threshold shown of 15 U/mL is the one used to determine seroconversion.

In our multivariable models, anti-SARS-CoV-2 antibody concentrations were lower in infliximab-compared with vedolizumab-treated patients in participants who received the BNT162b2 (fold change [FC] 0.29 [95% CI 0.21, 0.40], p<0.0001) and ChAdOx1 nCoV-19 [FC] 0.39 [95% CI 0.30, 0.51], p<0.0001) vaccines. Age ≥ 60 years, immunomodulator use, and current smoking were also independently associated with lower anti-SARS-CoV-2 antibody concentrations in participants who received either vaccine. Conversely, non-white ethnicity was associated with higher antibody concentrations following both vaccines (figure 2).

**Figure 2:**
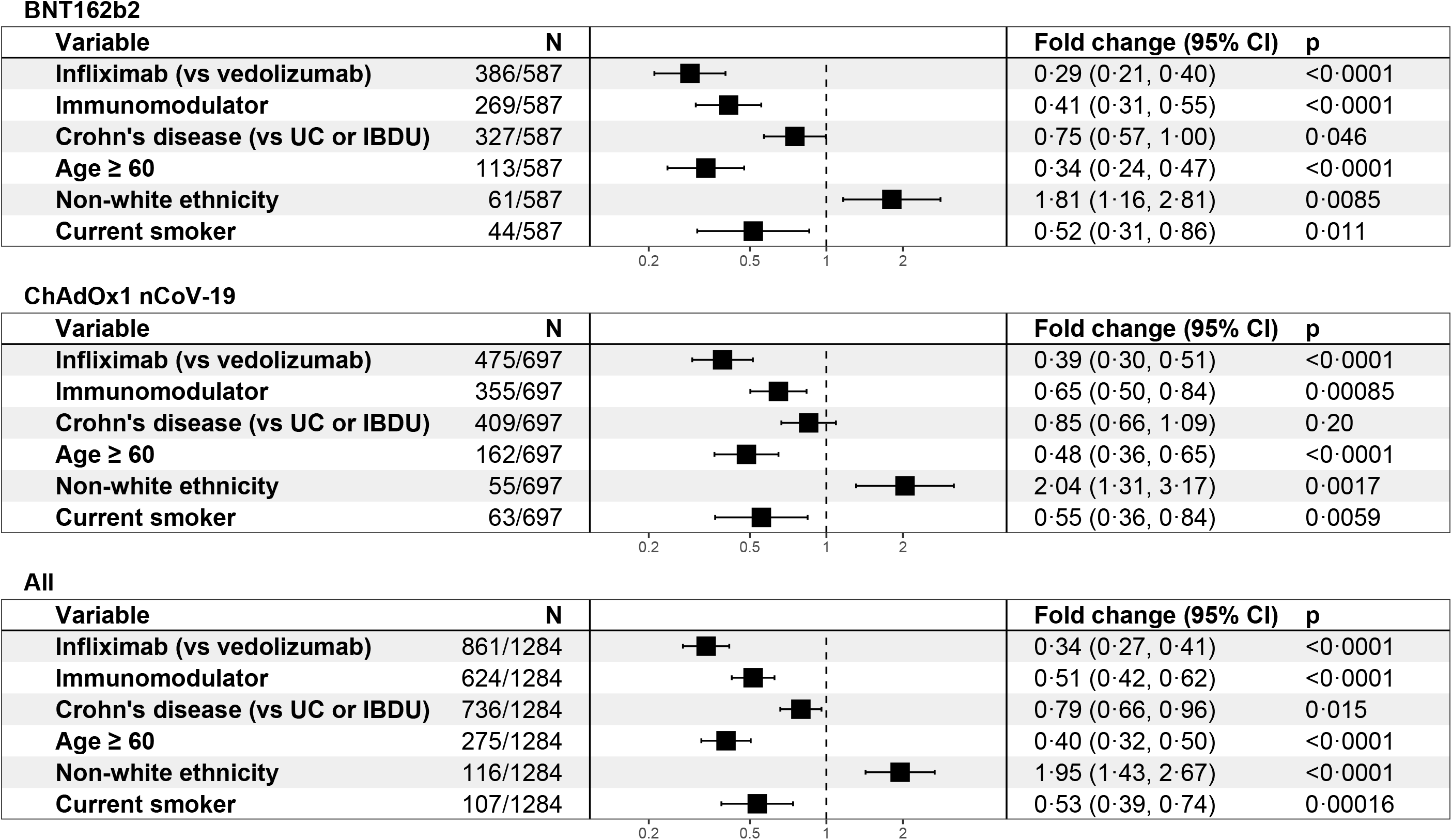
Exponentiated coefficients of linear regression models of log(anti-SARS-CoV-2 spike antibody concentration). The resultant values represent the fold change of antibody concentration associated with each variable. Each vaccine was modelled separately, and then a further model was created using all of the available data.

The 15-day rolling geometric mean of anti-SARS-CoV-2 antibody concentrations are shown in Figure 3. Three weeks after vaccination, we observed lower anti-SARS-CoV-2 (S) antibody concentrations in infliximab-compared to vedolizumab-treated patients following both vaccines. Sustained serological responses were observed in the vedolizumab-but not infliximab-treated patients.

**Figure 3:**
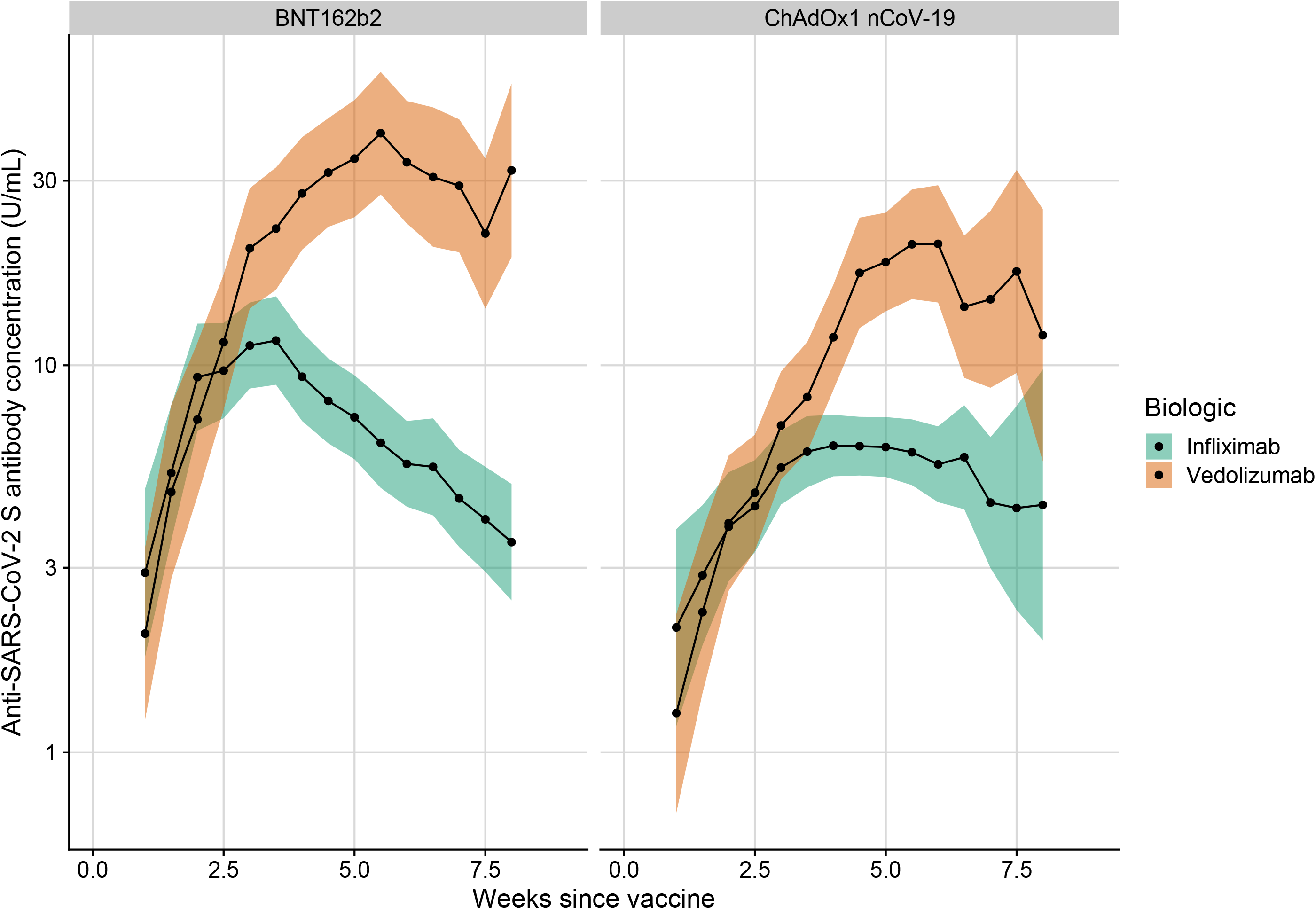
Rolling geometric mean antibody concentration over time stratified by biologic therapy (infliximab vs vedolizumab) and vaccine. Geometric means are calculated using a rolling 15 day window (i.e. 7 days either side of the day indicated). The shaded areas represent the 95% confidence intervals of the geometric means. Overall, data from 2126 participants (1427 on infliximab and 699 on vedolizumab) are included in this graph between 1 and 63 days post vaccination.

### Seroconversion following primary COVID-19 vaccination

The lowest rates of seroconversion were observed in participants treated with infliximab in combination with an immunomodulator with both the BNT162b2 (27.1%; 65/240) or ChAdOx1 nCoV-19 (20.2%; 60/297) vaccines. Highest rates of seroconversion were seen in patients treated with vedolizumab monotherapy who received the BNT162b2 (74.7%;124/166) or ChAdOx1 nCoV-19 (57.3%; 94/164) vaccines (Figure 4).

**Figure 4:**
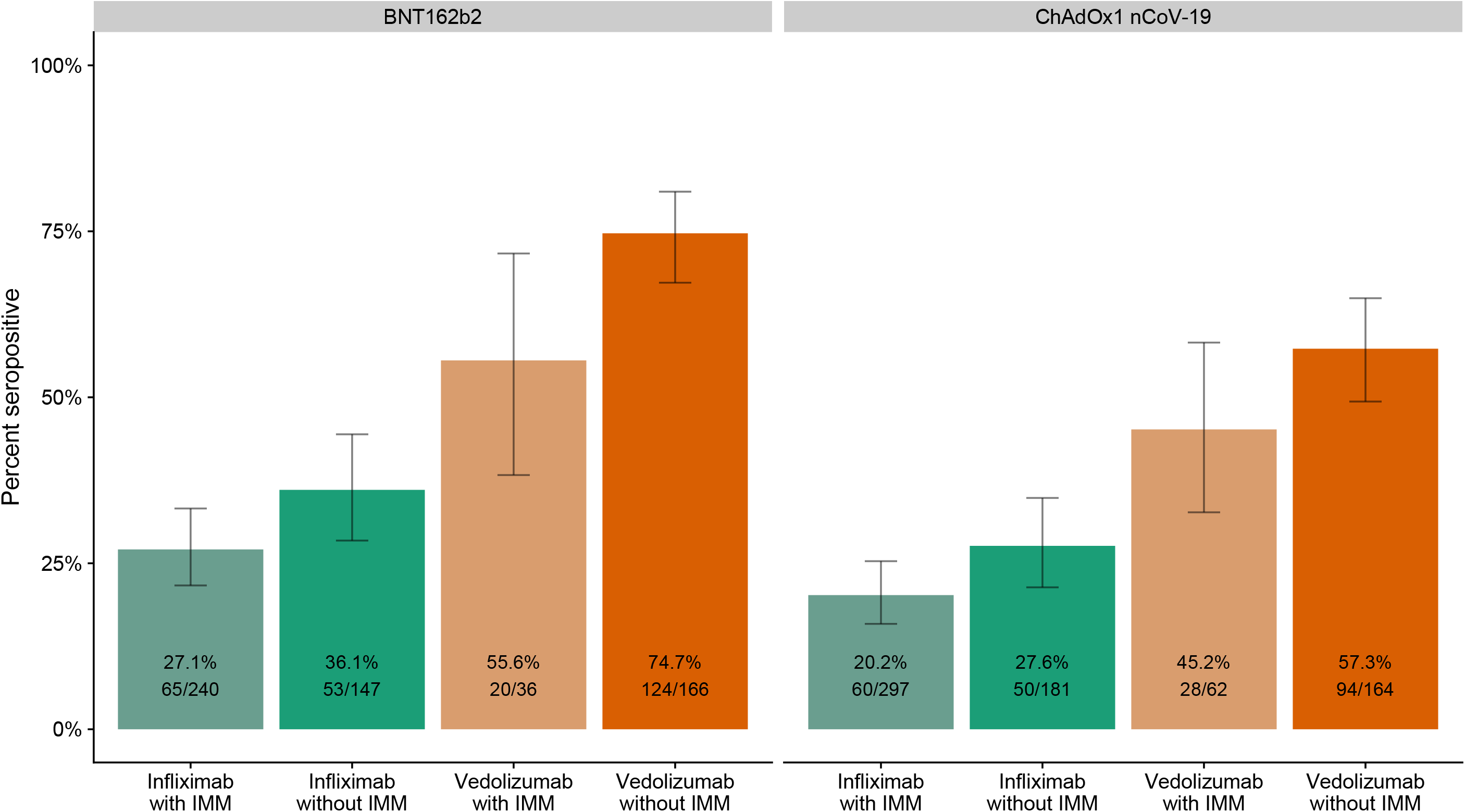
Percentages of participants with seroconversion defined by an anti-SARS-CoV-2 spike antibody concentration ≥ 15 U/mL, stratified by vaccine, biologic and immunomodulator use. Error bars represent the 95% confidence interval of the percentages. **Abbreviations**: IMM = immunomodulator

### Antibody responses following prior SARS-CoV-2 infection and two COVID-19 vaccine doses

Amongst participants with SARS-CoV-2 infection prior to vaccination, geometric mean [SD] anti-SARS-CoV-2 (S) antibody concentrations were lower in infliximab-compared with vedolizumab-treated patients in those who received a single-dose of BNT162b2 (191 U/mL [12.5] vs 1865 U/mL [8.0] P<0.0001) and ChAdOx1 nCoV-19 (185 U/mL [9.3] vs 752 [12.5] P=0.046) vaccines. In both infliximab- and vedolizumab-treated patients, antibody concentrations following vaccination were higher than those observed in patients without prior infection (Figure 5). Overall, across both vaccines, 82% (76/93) patients treated with infliximab and 97% (33/34) patients treated with vedolizumab seroconverted (p=0.041).

**Figure 5:**
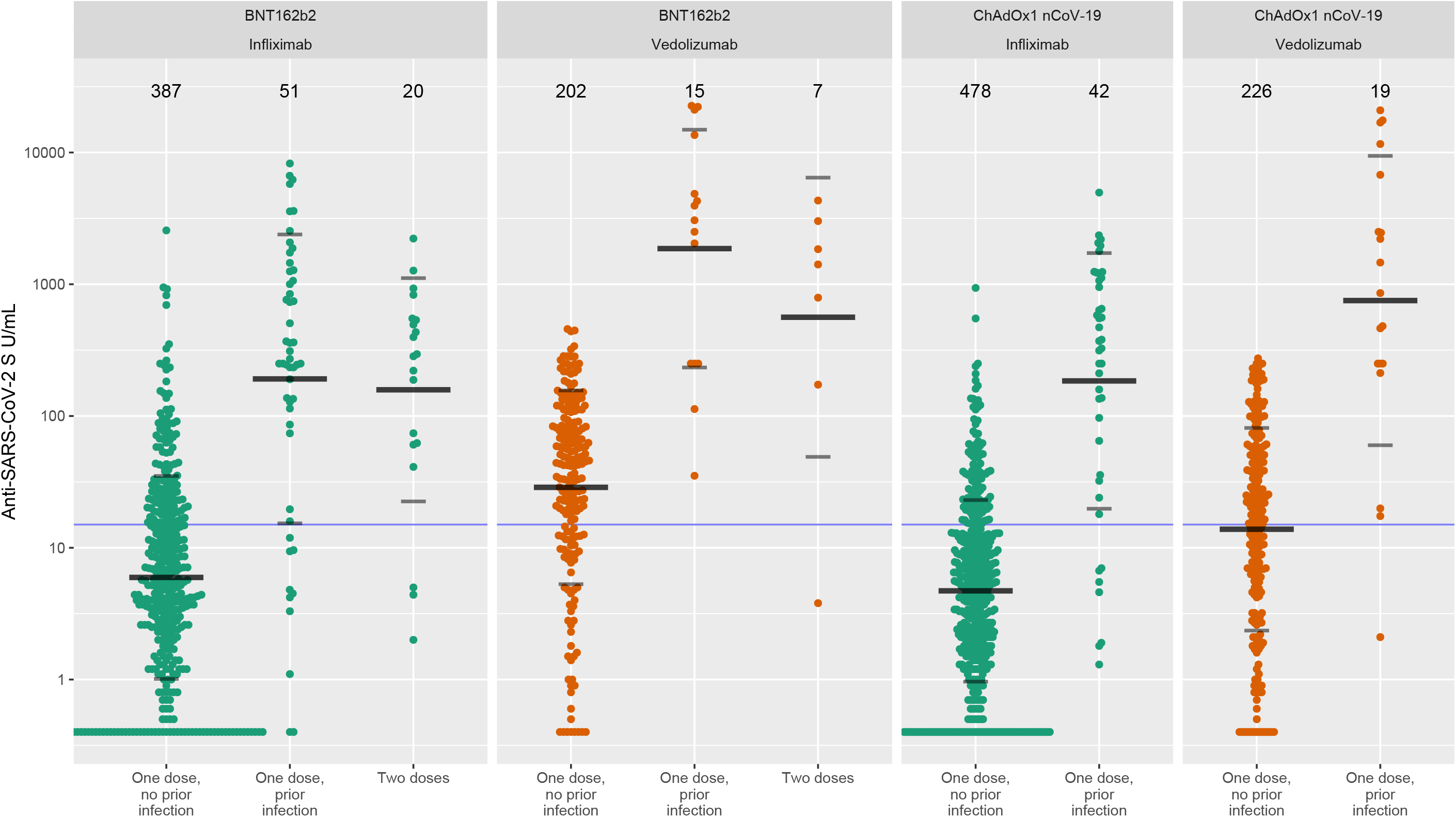
Anti-SARS-CoV-2 spike antibody concentration stratified by biologic therapy (infliximab vs vedolizumab), prior infection, number of doses and type of vaccine. The wider bar represents the geometric mean, while the narrower bars are drawn one geometric standard deviation either side of the geometric mean. The threshold shown of 15 U/mL is the one used to determine seroconversion.

Antibody responses were assessed in 27 patients following two doses of the BNT162b2 vaccine without serological evidence of prior infection (Figure 5). In both infliximab- and vedolizumab-treated patients, antibody levels and seroconversion rates were higher after two doses than after a primary vaccine without prior infection (geometric means infliximab 158 U/mL [7.0] vs 6.0 U/mL [5.9], p<0.0001; vedolizumab 562 U/mL [11.5] vs 28.8 U/mL [5.4], p = 0.018). After second-vaccine doses 85% (17/20) infliximab- and 86% (6/7) vedolizumab-treated patients seroconverted (p=0.68).

## Discussion

### Key results

We have shown that anti-SARS-CoV-2 spike antibody levels and rates of seroconversion are lower following vaccination with a single-dose of either BNT162b2 or ChAdOx1 nCoV-19 vaccines in patients with IBD treated with infliximab than vedolizumab. Combination therapy with an immunomodulator further attenuated immunogenicity to both vaccines in infliximab-treated patients. Reassuringly, however, a second exposure to antigen, either by vaccination after infection, or a second dose of vaccine led to seroconversion in most patients.

Direct comparisons between our data and the antibody responses reported in the vaccine registration trials are limited by differences in the assays used to define immunogenicity and the adoption of different thresholds to define seroconversion. No adequately powered studies have reported the effect of anti-TNF drugs on vaccine responses.^17^ Our findings are similar, however, to recent reports of the immunogenicity of the BNT162b2 and mRNA-1273 vaccines in transplant recipients and in patients with malignancy treated with anti-metabolite immunosuppression, conventional chemotherapy or immune checkpoint inhibitors.^18,19^ The authors showed fewer patients treated with potent immunosuppressants seroconverted than healthy controls. Importantly, as we have also shown here, second vaccine doses led to seroconversion in the cancer cohort. However, even after two antigen exposures, a small subset of patients (18% [20/113] infliximab-treated patients and 5% [2/41] vedolizumab-treated patients) in our study failed to mount an antibody response. To identify this group, and because the sustainability of antibody responses overall is unknown, serial measurement of antibody responses are indicated.

Urgent research is needed to understand the factors linked to non-response and how to potentiate long-term immunogenicity in this group. Strategies to be tested include the manipulation of timing of second vaccinations of second vaccinations, booster doses, the use of adjuvants and/or switching between vaccines with different mechanisms of action. Moreover, from the public health standpoint, recent case reports have shown that potent immunosuppression leads to chronic nasopharyngeal carriage and evolution of new SARS-CoV-2 variants.^20,21^ Whether this occurs in patients treated with anti-TNF therapy with impaired antibody response is an important conceptual concern.

Our data has other important findings relating to SARS-CoV-2 vaccine responses. We have demonstrated that antibody responses to SARS-CoV-2 vaccines are reduced in older individuals and current smokers. Smoking has also been associated with lower antibody responses to hepatitis B vaccination and faster decay of antibodies after vaccination with live attenuated and trivalent influenza vaccines.^22,23^ We have also demonstrated higher antibody responses to both the BNT162b2 and ChAdOx1 nCoV-19 vaccines in non-white participants. This might be explained by differences in genetics,^24^ gut microbiota,^25^ nutrition,^26^ and priming of the immune system by prior exposure to SARS-CoV-2 not detected by our pre-vaccination antibody test. Lower antibody concentrations were also observed in patients with Crohn’s disease when compared to patients with ulcerative colitis or IBD-unclassified. Despite evidence of defective mucosal immunity, previous vaccine studies involving patients with Crohn’s disease or ulcerative colitis have not shown attenuated antibody responses to vaccination in the absence of concomitant immunomodulator or biologic therapy.^6,7^

The cytokine TNF shapes multiple aspects of host immune responses, including T-cell dependent antibody production. Genetic ablation of TNF results in disruption of B-cell follicles in germinal centres with defective induction of antigen-induced antibody production.^27,28^ These biological properties may in part explain why TNF blockade is clinically beneficial in IMIDs, but also explain the increased risk of serious and opportunistic infections and impaired response to other vaccines.

Whilst our data are biologically plausible, we acknowledge the following limitations of our study. We have used an electrochemiluminescence immunoassay to measure antibody concentrations rather than using a neutralising assay. Although neutralisation assays are considered more biologically relevant, it is now established that anti receptor-binding domain antibodies, which target the spike protein component that engages host cells through ligation of angiotensin-converting enzyme 2, closely correlate with neutralisation assays.^29,30^ Second, we only assessed humoral responses to infection, and it is likely that protective immunity additionally requires induction of memory T cell responses. Finally, we investigated one anti-TNF drug, infliximab, only. However, we suspect that our key findings will apply to other anti-TNF biologics used to treat IMIDs, including adalimumab, certolizumab, golimumab, and etanercept. Further observational data will be required to elucidate the impact of other classes of therapies for IMIDs on SARS-CoV-2 vaccine immunogenicity.

Our findings have important implications for patients treated with anti-TNF drugs particularly those also treated with an immunomodulator. Poor antibody responses to a single-dose of vaccine unnecessarily exposes infliximab-treated patients to SARS-CoV-2 infection. However, because we observed higher rates of seroconversion in patients with two exposures to SARS-CoV-2 antigen, even in the presence of TNF blockade, these patients should be prioritised for optimally timed second doses. Until patients receive a second dose of vaccine they should consider that they are not protected from SARS-CoV-2 infection and continue to practice enhanced physical distancing and shielding if appropriate.

## Conclusion

Infliximab is associated with attenuated immunogenicity to a single-dose of the BNT162b2 and ChAdOx1 nCoV-19 SARS-CoV-2 vaccines in patients with inflammatory bowel disease. Immunomodulators further blunted immunogenicity rates to both vaccines. Reassuringly, vaccination after infection, or a second dose of vaccine led to seroconversion in most patients. Delayed second dosing should be avoided in patients treated with infliximab.

## Data Availability

The study protocol including the statistical analysis plan is available at www.clarityibd.org. Individual participant de-identified data that underlie the results reported in this article will be available immediately after publication for a period of 5 years. The data will be made available to investigators whose proposed use of the data has been approved by an independent review committee. Analyses will be restricted to the aims in the approved proposal. Proposals should be directed to. To gain access data requestors will need to sign a data access agreement.

## Contributions

NAK, JRG, CB, SS, NP, TA participated in the conception and design of this study. CB was the project manager and coordinated patient recruitment. RN and TJM coordinated all biochemical analyses and central laboratory aspects of the project. NAK, SL, JRG, NC, BH, DC, JRF, AF, PMI, NK, KBK, CAL, JM, SJM, RCGP, TR, PJS, AMV, TJM, SS, CWL, NP, TA were involved in the acquisition, analysis, or interpretation of data. Data analysis was done by NAK. Drafting of the manuscript was done by NAK, SL, JRG, NC, RN, DC, RCGP, SS, CWL, NP, TA. SS, NP and TA obtained the funding for the study. All the authors contributed to the critical review and final approval of the manuscript. NAK and TA have verified the underlying data.

## Declarations of interest

Dr. Kennedy reports grants from F. Hoffmann-La Roche AG, grants from Biogen Inc, grants from Celltrion Healthcare, grants from Galapagos NV, non-financial support from Immundiagnostik, during the conduct of the study; grants and non-financial support from AbbVie, grants and personal fees from Celltrion, personal fees and non-financial support from Janssen, personal fees from Takeda, personal fees and non-financial support from Dr Falk, outside the submitted work. Dr. Lin reports non-financial support from Pfizer, non-financial support from Ferring, outside the submitted work. Dr. Goodhand reports grants from F. Hoffmann-La Roche AG, grants from Biogen Inc, grants from Celltrion Healthcare, grants from Galapagos NV, non-financial support from Immundiagnostik, during the conduct of the study. Dr. Chee reports non-financial support from Ferring, personal fees and non-financial support from Pfizer, outside the submitted work. Dr. Cummings reports grants and personal fees from Samsung, Pfizer & Biogen; personal fees and non-financial support from Janssen & Abbvie; grants, personal fees and non-financial support from Takeda; personal fees from MSD, Sandoz, Celltrion & NAPP, outside the submitted work. Dr. Irving reports grants and personal fees from Takeda, grants from MSD, grants and personal fees from Pfizer, personal fees from Galapagos, personal fees from Gilead, personal fees from Abbvie, personal fees from Janssen, personal fees from Boehringer Ingelheim, personal fees from Topivert, personal fees from VH2, personal fees from Celgene, personal fees from Arena, personal fees from Samsung Bioepis, personal fees from Sandoz, personal fees from Procise, personal fees from Prometheus, outside the submitted work. Dr. Kamperidis reports personal fees from Janssen, outside the submitted work. Dr. Kok reports personal fees from Janssen, personal fees from Takeda, personal fees from PredictImmune, personal fees from Amgen, outside the submitted work. Dr. Lamb reports grants from Genentech, grants and personal fees from Janssen, grants and personal fees from Takeda, grants from AbbVie, personal fees from Ferring, grants from Eli Lilly, grants from Pfizer, grants from Roche, grants from UCB Biopharma, grants from Sanofi Aventis, grants from Biogen IDEC, grants from Orion OYJ, personal fees from Dr Falk Pharma, grants from AstraZeneca, outside the submitted work. Dr. Macdonald reports grants and personal fees from Takeda Pharmaceuticals, grants and personal fees from Biogen, personal fees and non-financial support from AbbVie, personal fees from Grifols, personal fees from Sandoz, personal fees from Celltrion, personal fees and non-financial support from Janssen, personal fees from Vifor Pharmaceuticals, personal fees from Predictimmune, personal fees from Bristol Myers Squibb, non-financial support from Ferring Pharmaceuticals, outside the submitted work. Prof. Pollok reports acting as consultant, advisory board member, speaker or recipient of educational grant from Dr Falk, Ferring, Janssen, Pharmacosmos and Takeda. Dr. Raine reports grants and personal fees from Abbvie, personal fees from BMS, personal fees from Celgene, personal fees from Ferring, personal fees from Gilead, personal fees from GSK, personal fees from LabGenius, personal fees from Janssen, personal fees from Mylan, personal fees from MSD, personal fees from Novartis, personal fees from Pfizer, personal fees from Sandoz, personal fees from Takeda, personal fees from Galapagos, personal fees from Arena, outside the submitted work. Dr Smith reports speaker fees and advisory board sponsorship from Janssen, Celltrion and Takeda outside the submitted work. Dr. Verma reports personal fees and non-financial support from Takeda, personal fees and non-financial support from Celltrion, personal fees and non-financial support from Merck Sharp & Dohme, outside the submitted work. Prof. Sebastian reports grants from Takeda, Abbvie, AMGEN, Tillots Pharma, personal fees from Jaansen, Takeda, Galapagos, Celltrion, Falk Pharma, Tillots pharma, Cellgene, Pfizer, Pharmacocosmos, outside the submitted work. Prof. Lees reports personal fees from Abbvie, personal fees from Janssen, personal fees from Pfizer, personal fees from Takeda, grants from Gilead, personal fees from Gilead, personal fees from Galapagos, personal fees from Iterative Scopes, personal fees from Trellus Health, personal fees from Celltion, personal fees from Ferring, personal fees from BMS, during the conduct of the study. Dr. Powell reports personal fees from Takeda, personal fees from Janssen, personal fees from Pfizer, personal fees from Bristol-Myers Squibb, personal fees from Abbvie, personal fees from Roche, personal fees from Lilly, personal fees from Allergan, personal fees from Celgene, outside the submitted work; and Dr. Powell has served as a speaker/advisory board member for Abbvie, Allergan, Bristol Myers Squibb, Celgene, Falk, Ferring, Janssen, Pfizer, Tillotts, Takeda and Vifor Pharma. Prof. Ahmad reports grants and non-financial support from F. Hoffmann-La Roche AG, grants from Biogen Inc, grants from Celltrion Healthcare, grants from Galapagos NV, non-financial support from Immundiagnostik, during the conduct of the study; personal fees from Biogen inc, grants and personal fees from Celltrion Healthcare, personal fees and non-financial support from Immundiagnostik, personal fees from Takeda, personal fees from ARENA, personal fees from Gilead, personal fees from Adcock Ingram Healthcare, personal fees from Pfizer, personal fees from Genentech, non-financial support from Tillotts, outside the submitted work. The following authors have nothing to declare: Neil Chanchlani, Ben Hamilton, Claire Bewshea, Rachel Nice, Shameer J Mehta, Timothy J McDonald.

## Acknowledgements

CLARITY IBD is a UK National Institute for Health Research (NIHR) Urgent Public Health Study. The NIHR Clinical Research Network supported study set-up, site identification, and delivery of this study. This was facilitated by Professor Mark Hull, the National speciality lead for Gastroenterology. We acknowledge the contribution of our Patient Advisory Group who helped shape the trial design around patient priorities. Our partners, Crohn’s and Colitis UK (CCUK), continue to support this group and participate in Study Management Team meetings. We thank Professor Danny Altmann, Professor Rosemary Boyton, Professor Graham Cooke and Dr Katrina Pollock for their helpful discussions and review of the data. Laboratory tests were undertaken by the Exeter Blood Sciences Laboratory at the Royal Devon and Exeter NHS Foundation Trust. The Exeter NIHR Clinical Research Facility coordinated sample storage and management. Tariq Malik and James Thomas from Public Health England, Guy Stevens, Katie Donelon, Elen de Lacy from Public Health Wales and Johanna Bruce from Public Health Scotland supported linkage of central SARS-CoV-2 PCR test results with study data. Roche Diagnostics Limited provided the Elecsys Anti-SARS-CoV-2 immunoassay for the study. SL is supported by a Wellcome GW4-CAT fellowship. NC acknowledges support from CCUK. CAL acknowledges support from the NIHR Newcastle Biomedical Research Centre and the support of the Programmed Investigation Unit at Royal Victoria Infirmary, Newcastle upon Tyne. TR acknowledges support with recruitment from the NIHR Cambridge BRC. CWL is funded by a UKRI Future Leaders Fellowship. NP is supported by the NIHR Imperial Biomedical Research Center (BRC). We acknowledge the study co-ordinators of the Exeter Inflammatory Bowel Disease Research Group: Marian Parkinson and Helen Gardner-Thorpe for their ongoing administrative support to the study. The sponsor of the study was the Royal Devon and Exeter NHS Foundation Trust.

## Patient involvement

We conducted an electronic survey to gauge the opinion of patients with IBD on the patient questionnaires to be delivered as part of the CLARITY IBD study. We surveyed 250 patients across 74 hospitals. All our proposed questions for study inclusion were rated as important or very important by at least 83% of participants. The Exeter IBD Patient Panel refined the questions included in the study questionnaire, reviewed the study protocol, supported the writing of the patient information sheets, and participated in testing of the electronic consent form and patient questionnaire. A member of the Exeter IBD Patient Panel sits on the study management committee, ensuring patient involvement in all aspects of study delivery, data analysis and dissemination of findings.

## Notes

### Clinical Trial

ISRCTN45176516

### Clinical Protocols

https://www.clarityibd.org

