## Supplemental files for "Infliximab is associated with attenuated immunogenicity to BNT162b2 and ChAdOx1 nCoV-19 SARS-CoV-2 vaccines"

### Table of Contents

Supplementary Table S1: Contributors to the CLARITY IBD study ....................2

Supplementary Table S2: Baseline characteristics of all participants in CLARITY IBD following primary vaccination against SARS-CoV-2, stratified by biologic....................18

Supplementary Table S3: Baseline characteristics of participants who had anti-SARS-CoV-2 spike antibodies measured 3 to 10 weeks following primary vaccination against SARS-CoV-2, stratified by vaccine administered...19

### Supplementary Table S1: Contributors to the CLARITY IBD study

| Affiliation | First name | Surname |
|---|---|---|
| Barts Health NHS Trust | Klaartje | Kok |
| | Farjhana | Bokth |
| | Bessie | Cipriano |
| | Caroline | Francia |
| | Nosheen | Khalid |
| | Hafiza | Khatun |
| | Ashley | Kingston |
| | Irish | Lee |
| | Anouk | Lehmann |
| | Kinnari | Naik |
| | Elise | Pabriaga |
| | Nicolene | Plaatjies |
| | Kevin | Samuels |
| Barts Health NHS Trust (paediatric) | Bessie | Cipriano |
| | Kevin | Samuels |
| | Nicolene | Plaatjies |
| | Hafiza | Khatun |
| | Farjana | Bokth |
| | Elise | Pabriaga |
| | Caroline | Francia |
| Basingstoke and North Hampshire Hospital | Rebecca | Saich |
| | Hayley | Cousins |
| | Wendy | Fraser |
| | Rachel | Thomas |
| | Matthew | Brown |

| Affiliation | First name | Surname |
|---|---|---|
| | Benjamin | White |
| Birmingham Women's and Children's NHS Foundation Trust | Rafeeq | Muhammed |
| | Rehana | Bi |
| | Catherine | Cotter |
| | Jayne | Grove |
| | Kate | Hong |
| | Ruth | Howman |
| | Monica | Mitchell |
| | Sugrah | Sultan |
| Bolton NHS Foundation Trust | Salil | Singh |
| | Chris | Dawe |
| | Robert | Hull |
| | Natalie | Silva |
| Borders General Hospital | Jonathan | Manning |
| | Lauren | Finlayson |
| Calderdale and Huddersfield NHS Foundation Trust | Sunil | Sonwalkar |
| | Naomi | Chambers |
| | Andrew | Haigh |
| | Lear | Matapure |
| Cambridge University Hospitals NHS Foundation Trust | Tim | Raine |
| | Varun | George |
| | Christina | Kapizioni |
| | Konstantina | Strongili |
| | Tina | Thompson |
| Chelsea and Westminster Hospital NHS Foundation Trust | Philip | Hendy |
| | Rhian | Bull |
| | Patricia | Costa |

| Affiliation | First name | Surname |
|---|---|---|
| | Lisa | Davey |
| | Hayley | Hannington |
| | Kribashnie | Nundlall |
| | Catarina | Martins |
| | Laura | Avanzi |
| | Jaime | Carungcong |
| | Sabrina | Barr |
| Chesterfield Royal Hospital | Kath | Phillis |
| | Rachel | Gascoyne |
| Countess Of Chester Hospital NHS Foundation Trust | Ian | London |
| | Jenny | Grounds |
| | Emmeline | Martin |
| | Susie | Pajak |
| Darlington Memorial Hospital | Anjan | Dhar |
| | Ellen | Brown |
| | Amanda | Cowton |
| | Kimberley | Stamp |
| Dartford and Gravesham NHS Trust | Ben | Warner |
| | Carmel | Stuart |
| | Louise | Lacey |
| The Dudley Group NHS Foundation Trust | Shanika | de Silva |
| | Clare | Allcock |
| | Philip | Harvey |
| East and North Hertfordshire NHS Trust | Johanne | Brooks |
| | Pearl | Baker |
| | Hannah | Beadle |
| | Carina | Cruz |

| Affiliation | First name | Surname |
|---|---|---|
| | Debbie | Potter |
| East Lancashire Hospitals NHS Trust | Joe | Collum |
| | Farzana | Masters |
| East Suffolk and North Essex NHS Foundation Trust | Achuth | Shenoy |
| | Alison | O'Kelly |
| Glangwili Hospital | Aashish | Kumar |
| | Samantha | Coetzee |
| | Mihaela | Peiu |
| Great Ormond Street Hospital | Edward | Gaynor |
| | Sibongile | Chadokufa |
| | Bonita | Huggett |
| | Hamza | Meghari |
| | Sara | El-Khouly |
| | Fevronia | Kiparissi |
| | Waffa | Girshab |
| Great Western Hospitals NHS Foundation Trust | Andrew | Claridge |
| | Emily | Fowler |
| | Laura | McCafferty |
| Guy's and St Thomas' NHS Foundation Trust | Peter | Irving |
| | Karolina | Christodoulides |
| | Angela | Clifford |
| | Patrick | Dawson |
| | Sailish | Honap |
| | Samuel | Lim |
| | Raphael | Luber |
| | Karina | Mahiouz |
| | Susanna | Meade |

| Affiliation | First name | Surname |
|---|---|---|
| | Parizade | Raymode |
| | Rebecca | Reynolds |
| | Anna | Stanton |
| | Sherill | Tripoli |
| | Naomi | Hare |
| The Hillingdon Hospitals NHS Foundation Trust | Yih Harn | Siaw |
| | Lane | Manzano |
| | Jonathan | Segal |
| | Ibrahim | Al-Bakir |
| | Imran | Khakoo |
| Homerton University Hospital Foundation Trust | Nora | Thoua |
| | Katherine | Davidson |
| | Jagrul | Miah |
| | Alex | Hall |
| Hull University Teaching Hospitals NHS Trust | Shaji | Sebastian |
| | Melony | Hayes |
| | Sally | Myers |
| | Alison | Talbot |
| | Jack | Turnbull |
| | Emma | Whitehead |
| | Katie | Stamp |
| | Alison | Pattinson |
| | Verghese | Mathew |
| | Leanne | Sherris |
| Imperial College Healthcare NHS Trust | Lucy | Hicks |
| | Tara-Marie | Byrne |
| | Leilani | Cabreros |

| Affiliation | First name | Surname |
|---|---|---|
| | Hannah | Downing-Wood |
| | Sophie | Hunter |
| | Mohammad Aamir | Saifuddin |
| | Hemanth | Prabhudev |
| | Sharmili | Balarajah |
| James Paget University Hospitals NHS Foundation Trust | Helen | Sutherland |
| Kettering General Hospital | Ajay M | Verma |
| | Juliemol | Sebastian |
| | Mohammad Farhad | Peerally |
| King's College Hospital NHS Foundation Trust | Alexandra | Kent |
| | Lee Meng | Choong |
| | Benedetta | Pantaloni |
| | Pantelis | Ravdas |
| King's College Hospital NHS Foundation Trust (paediatric) | Babu | Vadamalayan |
| King’s Mill Hospital | Stephen | Foley |
| | Becky | Arnold |
| | Cheryl | Heeley |
| | Wayne | Lovegrove |
| Liverpool University Hospitals NHS Foundation Trust | Philip J | Smith |
| | Giovanna | Bretland |
| | Sarah | King |
| | Martina | Lofthouse |
| | Lindsey | Rigby |
| | Sreedhar | Subramanian |
| | David | Tyrer |
| | Kate | Martin |

| Affiliation | First name | Surname |
|---|---|---|
| | Christopher | Probert |
| London North West University Healthcare NHS Trust | Nikolaos | Kamperidis |
| | Temi | Adedoyin |
| | Manisha | Baden |
| | Jeannette | Brown |
| | Feba | Chacko |
| | Michela | Cicchetti |
| | Mohammad | Saifuddin |
| | Priya | Yesupatham |
| Maidstone and Tunbridge Wells NHS Trust | Rohit | Gowda |
| | Maureen | Williams |
| Manchester University NHS Foundation Trust | Karen | Kemp |
| | Rima | Akhand |
| | Glaxy | Gray |
| | Anu | John |
| | Maya | John |
| | Diamond | Sathe |
| | Jennifer | Soren |
| The Mid Yorkshire Hospitals NHS Trust | Michael | Sprakes |
| | Julie | Burton |
| | Patricia | Kane |
| | Stephanie | Lupton |
| Milton Keynes University Hospital | George | MacFaul |
| | Diane | Scaletta |
| | Loria | Siamia |
| | Felicity | Williams |
| Newcastle Hospitals NHS | Chris | Lamb |

| Affiliation | First name | Surname |
|---|---|---|
| Foundation Trust | Mary | Doona |
| | Ashleigh | Hogg |
| | Lesley | Jeffrey |
| | Andrew | King |
| | R Alexander | Speight |
| Ninewells Hospital & Medical School | Craig | Mowat |
| | Debbie | Rice |
| | Susan | MacFarlane |
| | Anne | MacLeod |
| | Samera | Mohammed |
| Norfolk and Norwich University Hospitals NHS Foundation Trust | Mary | Anne Morris |
| | Louise | Coke |
| | Grace | Hindle |
| | Eirini | Kolokouri |
| | Catherine | Wright |
| North Bristol NHS Trust | Melanie | Lockett |
| | Charlotte | Cranfield |
| | Louise | Jennings |
| | Ankur | Srivastava |
| | Lana | Ward |
| | Nouf | Jeynes |
| North Tyneside General Hospital | Praveen | Rajasekhar |
| | Lisa | Gallagher |
| | Linda | Patterson |
| | Jill | Ward |
| | Rae | Basnett |
| | Judy | Murphy |

| Affiliation | First name | Surname |
|---|---|---|
| | Lauren | Parking |
| | Emma | Lawson |
| Nottingham University Hospitals NHS Trust | David | Devadason |
| | Gordon | Moran |
| | Neelam | Khan |
| | Lauren | Tarr |
| The Pennine Acute Hospitals NHS Trust | Jimmy | Limdi |
| | Kay | Goulden |
| | Asad | Javed |
| | Lauren | McKenzie |
| Portsmouth Hospitals NHS Trust | Pradeep | Bhandari |
| | Michelle | Baker-Moffatt |
| | Joanne | Dash |
| The Queen Elizabeth Hospital Kings Lynn NHS Trust | Alan | Wiles |
| | Hannah | Bloxham |
| | Jose | Dias |
| | Ellie | Graham |
| Queen Elizabeth University Hospital, Glasgow | Jonathan | Macdonald |
| | Shona | Finan |
| | Faye | McMeeken |
| | Stephanie | Shields |
| | John Paul | Seenan |
| Royal Berkshire NHS Foundation Trust | Des | DeSilva |
| | Ofori | Boateng |
| | Holly | Lawrence |
| | Susanna | Malkakorpi |
| The Royal Bournemouth and | Simon | Whiteoak |

| Affiliation | First name | Surname |
|---|---|---|
| Christchurch Hospitals NHS Foundation Trust | Kelli | Edger-Earley |
| Royal Cornwall Hospitals NHS Trust | Sarah | Ingram |
| | Sharon | Botfield |
| | Fiona | Hammonds |
| | Clare | James |
| Royal Devon and Exeter NHS Foundation Trust | Tariq | Ahmad |
| | Gemma | Aspinall |
| | Sarah | Hawkins |
| | Suzie | Marriott |
| | Clare | Redstone |
| | Halina | Windak |
| Royal Free London NHS Foundation Trust | Charles | Murray |
| | Cynthia | Diaba |
| | Fexy | Joseph |
| | Glykeria | Pakou |
| Royal Glamorgan Hospital | James | Berrill |
| | Natalie | Stroud |
| | Carla | Pothecary |
| | Lisa | Roche |
| | Keri | Turner |
| | Lisa | Deering |
| | Lynda | Israel |
| Royal Gwent Hospital | Evelyn | Baker |
| | Sean | Cutler |
| | Rina | Mardania Evans |
| | Maxine | Nash |

| Affiliation | First name | Surname |
|---|---|---|
| Royal Hampshire County Hospital | John | Gordon |
| | Emma | Levell |
| | Silvia | Zagalo |
| Royal Hospital for Sick Children, Edinburgh | Richard | Russell |
| | Paul | Henderson |
| | Margaret | Millar |
| Royal Manchester Children's Hospital | Andrew | Fagbemi |
| | Felicia | Jennings |
| | Imelda | Mayor |
| | Jill | Wilson |
| Royal Surrey County Hospital | Christopher | Alexakis |
| | Natalia | Michalak |
| Royal United Hospitals Bath | John | Saunders |
| | Helen | Burton |
| | Vanessa | Cambridge |
| | Tonia | Clark |
| | Charlotte | Ekblad |
| | Sarah | Hierons |
| | Joyce | Katebe |
| | Emma | Saunsbury |
| | Rachel | Perry |
| The Royal Wolverhampton NHS Trust | Matthew | Brookes |
| | Kathryn | Davies |
| | Marie | Green |
| | Ann | Plumbe |
| Salford Royal NHS Foundation Trust | Clare | Ormerod |
| | Helen | Christensen |

| Affiliation | First name | Surname |
|---|---|---|
| | Anne | Keen |
| | Jonathan | Ogor |
| Salisbury District Hospital | Alpha | Anthony |
| | Emily | Newitt |
| Sandwell and West Birmingham NHS Trust | Edward | Fogden |
| | Kalisha | Russell |
| Sheffield Teaching Hospitals NHS Foundation Trust | Anne | Phillips |
| | Muaad | Abdulla |
| Shrewsbury and Telford Hospital NHS Trust | Jeff | Butterworth |
| | Colene | Adams |
| | Elizabeth | Buckingham |
| | Danielle | Childs |
| | Alison | Magness |
| | Jo | Stickley |
| Singleton Hospital | Caradog | Thomas |
| | Elaine | Brinkworth |
| | Lynda | Connor |
| | Amanda | Cook |
| | Tabitha | Rees |
| Somerset NHS Foundation Trust | Emma | Wesley |
| | Alison | Moss |
| South Tees Hospitals NHS Foundation Trust | Arvind | Ramadas |
| | Julie | Tregonning |
| Southend University Hospital NHS Foundation Trust | Ioannis | Koumoutsos |
| | Viji | George |
| | Swapna | Kunhunny |
| | Sophie | Laverick |

| Affiliation | First name | Surname |
|---|---|---|
| St George's University Hospitals NHS Foundation Trust | Kamal | Patel |
| | Mariam | Ali |
| | Hilda | Mhandu |
| | Aleem | Rana |
| | Katherine | Spears |
| | Joana | Teixeira |
| | Richard | Pollok |
| | Mark | Mencias |
| | Abigail | Seaward |
| | Jessica | Sousa |
| St George's University Hospitals NHS Foundation Trust (paediatric) | Nicholas | Reps |
| | Rebecca | Martin |
| St James's University Hospital | Christian | Selinger |
| | Jenelyn | Carbonell |
| | Felicia | Onovira |
| | Doris | Quartey |
| Stockport NHS Foundation Trust | Zahid | Mahmood |
| | Racheal | Campbell |
| | Liane | Marsh |
| Surrey and Sussex Healthcare NHS Trust | Monira | Rahman |
| | Sarah | Davies |
| | Ruth | Habibi |
| | Ellen | Jessup-Dunton |
| | Teishel | Joefield |
| | Reina | Layug |
| Tameside and Glossop Integrated Care NHS Foundation Trust | Vinod | Patel |
| | Joanne | Vere |

| Affiliation | First name | Surname |
|---|---|---|
| Torbay and South Devon NHS Foundation Trust | Gareth | Walker |
| | Stacey | Atkins |
| | Jasmine | Growdon |
| | Charlotte | McNeill |
| University Hospitals Birmingham NHS Foundation Trust | Rachel | Cooney |
| | Lillie | Bennett |
| | Louise | Bowlas |
| | Sharafaath | Shariff |
| University Hospitals Bristol NHS Foundation Trust | Aileen | Fraser |
| | Katherine | Belfield |
| University Hospitals of Derby and Burton NHS Foundation Trust | Said | Din |
| | Catherine | Addleton |
| | Marie | Appleby |
| | Johanna | Brown |
| | Kathleen | Holding |
| University Hospitals of Leicester NHS Trust | John | deCaestecker |
| | Olivia | Watchorn |
| University Hospitals Plymouth NHS Trust | Chris | Hayward |
| | Susan | Inniss |
| | Lucy | Pritchard |
| United Lincolnshire Hospitals NHS Trust | Jervoise | Andreyev |
| | Caroline | Hayhurst |
| | Carol | Lockwood |
| | Lynn | Osborne |
| | Amanda | Roper |
| | Karen | Warner |
| | Julia | Hindle |

| Affiliation | First name | Surname |
|---|---|---|
| University College London Hospitals NHS Foundation Trust | Shameer | Mehta |
| | James | Bell |
| | William | Blad |
| | Lisa | Whitley |
| University Hospital Llandough | Durai | Dhamaraj |
| | Mark | Baker |
| University Hospital Southampton NHS Foundation Trust | Fraser | Cummings |
| | Clare | Harris |
| | Amy | Jones |
| | Liga | Krauze |
| | Sohail | Rahmany |
| | Audrey | Torokwa |
| University Hospital of Wales (paediatric) | Amar | Wahid |
| | Zoe | Morrison |
| West Hertfordshire Hospitals NHS Trust | Rakesh | Chaudhary |
| | Melanie | Claridge |
| | Chiara | Ellis |
| | Cheryl | Kemp |
| | Ogwa | Tobi |
| West Middlesex University Hospital | Emma | Johnston |
| | Metod | Oblak |
| | Richard | Appleby |
| West Suffolk NHS Foundation Trust | Marium | Asghar |
| Western General Hospital | Charlie | Lees |
| | Debbie | Alexander |
| | Kate | Covil |

| Affiliation | First name | Surname |
|---|---|---|
| | Lauranne | Derikx |
| | Sryros | Siakavellas |
| | Helen | Baxter |
| | Scott | Robertson |
| Withybush General Hospital | Kerrie | Johns |
| | Rachel | Hughes |
| | Janet | Phipps |
| | Abigail | Taylor |
| Yeovil District Hospital NHS Foundation Trust | Katie | Smith |
| | Linda | Howard |
| | Dianne | Wood |
| York Teaching Hospital NHS Foundation Trust | Ajay | Muddu |
| | Laura | Barman |
| | Janine | Mallinson |
| Ysbyty Gwynedd | Iona | Thomas |
| | Kelly | Andrews |
| | Caroline | Mulvaney Jones |
| | Julia | Roberts |

### Supplementary Table S2: Baseline characteristics of all participants in CLARITY IBD following primary vaccination against SARS-CoV-2, stratified by biologic

| Variable | | Infliximab | Vedolizumab | Overall | p |
|---|---|---|---|---|---|
| Vaccine | BNT162b2 | 46·7% (741/1587) | 45·1% (342/758) | 46·2% (1083/2345) | 0·48 |
| | ChAdOx1 nCoV-19 | 53·3% (846/1587) | 54·9% (416/758) | 53·8% (1262/2345) | |
| Age (years) | | 41·2 (31·0 - 54·4) | 51·0 (37·0 - 63·8) | 43·8 (32·4 - 57·4) | <0·0001 |
| Sex | Female | 48·6% (770/1585) | 48·6% (367/755) | 48·6% (1137/2340) | 0·47 |
| | Male | 51·4% (814/1585) | 51·1% (386/755) | 51·3% (1200/2340) | |
| | Intersex | 0·0% (0/1585) | 0·0% (0/755) | 0·0% (0/2340) | |
| | Prefer not to say | 0·1% (1/1585) | 0·3% (2/755) | 0·1% (3/2340) | |
| Ethnicity | White | 92·3% (1461/1583) | 90·3% (679/752) | 91·6% (2140/2335) | 0·38 |
| | Asian | 4·9% (77/1583) | 6·5% (49/752) | 5·4% (126/2335) | |
| | Mixed | 1·6% (26/1583) | 1·9% (14/752) | 1·7% (40/2335) | |
| | Black | 0·8% (13/1583) | 0·7% (5/752) | 0·8% (18/2335) | |
| | Other | 0·4% (6/1583) | 0·7% (5/752) | 0·5% (11/2335) | |
| Diagnosis | Crohn's disease | 63·7% (1011/1587) | 38·7% (293/758) | 55·6% (1304/2345) | <0·0001 |
| | Ulcerative colitis or IBD-unclassified | 36·3% (576/1587) | 61·3% (465/758) | 44·4% (1041/2345) | |
| Duration of IBD (years) | | 8·0 (4·0 - 16·0) | 10·0 (4·0 - 18·0) | 9·0 (4·0 - 16·0) | 0·00035 |
| Age at IBD diagnosis (years) | | 28·6 (20·9 - 41·7) | 35·3 (24·2 - 49·2) | 30·3 (21·6 - 44·1) | <0·0001 |
| Immunomodulators at vaccine | | 61·0% (968/1586) | 21·4% (162/756) | 48·2% (1130/2342) | <0·0001 |
| 5-ASA | | 22·8% (362/1586) | 32·7% (247/756) | 26·0% (609/2342) | <0·0001 |
| Steroids | | 3·3% (52/1586) | 7·7% (58/756) | 4·7% (110/2342) | <0·0001 |
| BMI (kg/m$^2$) | | 25·9 (22·8 - 30·3) | 26·0 (23·1 - 30·2) | 26·0 (22·9 - 30·2) | 0·72 |
| Heart disease | | 3·0% (48/1587) | 6·2% (47/758) | 4·1% (95/2345) | 0·00046 |
| Diabetes | | 3·5% (55/1587) | 8·3% (63/758) | 5·0% (118/2345) | <0·0001 |
| Lung disease | | 13·7% (218/1587) | 18·2% (138/758) | 15·2% (356/2345) | 0·0056 |
| Kidney disease | | 1·1% (18/1587) | 2·2% (17/758) | 1·5% (35/2345) | 0·045 |
| Cancer | | 0·3% (5/1587) | 2·2% (17/758) | 0·9% (22/2345) | <0·0001 |
| Smoker | Yes | 9·7% (153/1580) | 7·3% (55/753) | 8·9% (208/2333) | 0·0030 |
| | Not currently | 32·8% (519/1580) | 39·6% (298/753) | 35·0% (817/2333) | |
| | Never | 57·5% (908/1580) | 53·1% (400/753) | 56·1% (1308/2333) | |
| Exposure to documented cases of COVID-19 | | 10·2% (161/1581) | 9·3% (70/754) | 9·9% (231/2335) | 0·55 |
| Income deprivation score | | 0·092 (0·054 - 0·153) | 0·088 (0·055 - 0·146) | 0·091 (0·054 - 0·151) | 0·72 |
| Active disease (PRO2) | | 6·1% (92/1513) | 12·4% (89/718) | 8·1% (181/2231) | <0·0001 |

Table includes participants with antibody levels measured outside of the week 3-10 window as well as those with prior infection or with two doses of vaccine; this represents all participants included in figures 1-5 of the main paper.

**Abbreviations:** IBD = inflammatory bowel disease; 5-ASA = 5-aminosalicylic acid; BMI = Body Mass Index; PRO2 = IBD disease activity.

Values presented are median (interquartile range) or percentage (numerator/denominator). P values represent the results of a Mann Whitney U, Kruskal Wallis or Fisher’s exact test.

### Supplementary Table S3: Baseline characteristics of participants who had anti-SARS-CoV-2 spike antibodies measured 3 to 10 weeks following primary vaccination against SARS-CoV-2, stratified by vaccine administered

| Variable | | BNT162b2 | ChAdOx1 nCoV-19 | Overall | p |
|---|---|---|---|---|---|
| Biologic | Vedolizumab | 34·3% (202/589) | 32·1% (226/704) | 33·1% (428/1293) | 0·41 |
| | Infliximab | 65·7% (387/589) | 67·9% (478/704) | 66·9% (865/1293) | |
| Age (years) | | 43·9 (32·1 - 56·3) | 43·8 (33·3 - 58·9) | 43·8 (32·8 - 57·6) | 0·51 |
| Sex | Female | 48·4% (285/589) | 49·9% (349/699) | 49·2% (634/1288) | 0·52 |
| | Male | 51·4% (303/589) | 50·1% (350/699) | 50·7% (653/1288) | |
| | Intersex | 0·0% (0/589) | 0·0% (0/699) | 0·0% (0/1288) | |
| | Prefer not to say | 0·2% (1/589) | 0·0% (0/699) | 0·1% (1/1288) | |
| Ethnicity | White | 90·0% (530/589) | 92·1% (642/697) | 91·1% (1172/1286) | 0·65 |
| | Asian | 6·6% (39/589) | 5·6% (39/697) | 6·1% (78/1286) | |
| | Mixed | 2·4% (14/589) | 1·4% (10/697) | 1·9% (24/1286) | |
| | Black | 0·7% (4/589) | 0·6% (4/697) | 0·6% (8/1286) | |
| | Other | 0·3% (2/589) | 0·3% (2/697) | 0·3% (4/1286) | |
| Diagnosis | Crohn's disease | 55·7% (328/589) | 58·5% (412/704) | 57·2% (740/1293) | 0·31 |
| | Ulcerative colitis or IBD-unclassified | 44·3% (261/589) | 41·5% (292/704) | 42·8% (553/1293) | |
| Duration of IBD (years) | | 9·0 (4·0 - 16·0) | 9·0 (4·0 - 17·0) | 9·0 (4·0 - 16·0) | 0·38 |
| Age at IBD diagnosis (years) | | 30·0 (21·9 - 42·6) | 30·6 (21·9 - 44·5) | 30·3 (21·9 - 43·7) | 0·60 |
| Immunomodulators at vaccine | | 45·9% (270/588) | 50·7% (357/704) | 48·5% (627/1292) | 0·094 |
| 5-ASA | | 25·0% (147/588) | 26·6% (187/704) | 25·9% (334/1292) | 0·57 |
| Steroids | | 4·6% (27/588) | 5·0% (35/704) | 4·8% (62/1292) | 0·79 |
| BMI (kg/m$^2$) | | 25·6 (22·7 - 30·2) | 26·1 (23·1 - 30·6) | 26·0 (22·9 - 30·4) | 0·21 |
| Heart disease | | 5·4% (32/589) | 3·8% (27/704) | 4·6% (59/1293) | 0·18 |
| Diabetes | | 4·9% (29/589) | 5·1% (36/704) | 5·0% (65/1293) | 0·90 |
| Lung disease | | 15·1% (89/589) | 15·1% (106/704) | 15·1% (195/1293) | 1·0 |
| Kidney disease | | 1·5% (9/589) | 1·4% (10/704) | 1·5% (19/1293) | 1·0 |
| Cancer | | 1·0% (6/589) | 1·0% (7/704) | 1·0% (13/1293) | 1·0 |
| Smoker | Yes | 7·5% (44/588) | 9·0% (63/699) | 8·3% (107/1287) | 0·042 |
| | Not currently | 32·3% (190/588) | 37·6% (263/699) | 35·2% (453/1287) | |
| | Never | 60·2% (354/588) | 53·4% (373/699) | 56·5% (727/1287) | |
| Exposure to documented cases of COVID-19 | | 11·2% (66/588) | 7·4% (52/699) | 9·2% (118/1287) | 0·020 |
| Income deprivation score | | 0·081 (0·049 - 0·141) | 0·090 (0·054 - 0·151) | 0·086 (0·052 - 0·147) | 0·057 |
| Active disease (PRO2) | | 7·8% (44/567) | 6·4% (43/669) | 7·0% (87/1236) | 0·37 |

**Abbreviations:** IBD = inflammatory bowel disease; 5-ASA = 5-aminosalicylic acid; BMI = Body Mass Index; PRO2 = IBD disease activity.

Values presented are median (interquartile range) or percentage (numerator/denominator). P values represent the results of a Mann Whitney U, Kruskal Wallis or Fisher's exact test.
